# Reproductive Ageing in Women (RAW) Questionnaire: multi-phase development and validation of a questionnaire for the classification of menopause stage

**DOI:** 10.64898/2026.03.25.26349141

**Authors:** Laura E Pernoud, Paul A Gardiner, Melinda M Dean, Jamie L Noll, Mia A Schaumberg

## Abstract

**Objectives:** Accurate classification of menopausal stage is fundamental to midlife health research. Although the Stages of Reproductive Ageing Workshop (STRAW+10) criteria provide gold-standard criteria, their application in research settings is inconsistent. Classification challenges are compounded in individuals without observable menstrual cycles due to surgical or contraceptive-induced amenorrhoea. The Reproductive Ageing in Women (RAW) Questionnaire and accompanying classification Framework was developed and validated to improve consistency and inclusivity when classifying menopausal stage.

**Study design:** A multi-phase study was conducted between May 2022 and July 2025. Phase one involved questionnaire development based on STRAW+10 and the Menopause-Specific Quality of Life Questionnaire. Phase two assessed content validity via expert review (n=3). Phase three evaluated face validity using think-aloud interviews and focus groups (n=14). Phase four validated RAW within a cross-sectional cohort study (n=156), and assessed construct validity (n=30), test–retest reliability (6-21 days; n=128; Kendall’s Tau-b and Cohen’s kappa), and biological validity using follicle stimulating hormone (FSH).

**Results:** Feedback supported clarity, relevance and usability, with refinements improving inclusivity for surgical and contraceptive-induced amenorrhoea. Construct validity demonstrated consistent application of classification criteria. Questionnaire classification showed 93% concordance with self-identified menopausal status in the construct sample and 87.8% agreement within the cohort sample. Test–retest reliability was excellent (τb=0.940, p<0.001). Follicle stimulating hormone levels differed across RAW-classified stages (p<0.001), with 96.1% concordance between RAW pre and postmenopausal classifications and FSH thresholds.

**Conclusions:** Prioritising menstrual characteristics while incorporating age and symptom criteria improves methodological consistency and inclusivity in menopausal stage classification. Longitudinal validation is warranted to assess temporal sensitivity across the menopausal transition.

## 1. Background

The menopausal transition is characterised by progressive change in reproductive hormone dynamics and menstrual cycle characteristics, which reflect a decline in ovarian reserve (1, 2). Although menopause itself is defined retrospectively as 12 months of amenorrhea following the final menstrual period (FMP), the years preceding encompass substantial hormonal fluctuations and physiological adaptation (1, 3). These changes have important implications for cardiometabolic, skeletal, inflammatory and neurocognitive health, positioning midlife as a critical window for disease risk modification in women (4).

To facilitate classification of reproductive ageing, the Stages of Reproductive Ageing Workshop (STRAW+10) provide gold-standard guidelines that define early, peak and late reproductive stages, and the early and late menopausal transition (5). Each stage is defined by specific primary criteria, based on menstrual cycle characteristics, and supportive criteria, including hormone markers and vasomotor symptoms. Despite widespread recognition of these criteria, application in research settings remains inconsistent (6, 7). In some studies, menopausal classification is inferred from single time-point hormone levels without adherence to menstrual cycle characteristics (8). In others, simplified classifications such as reporting at least one menstrual cycle within the preceding 12 months are used to define premenopause (9), potentially obscuring distinctions between late reproductive stage and the menopausal transition as defined by STRAW+10 (5).

Inconsistent classification reduces comparability across studies and may attenuate stage-specific associations. Given the well-established influence of reproductive hormones on metabolic, inflammatory and vascular physiology, misclassification may have important implications for interpretation of midlife health outcomes (4, 10, 11). Classification challenges are further compounded in individuals for whom menstrual cycle characteristics cannot be determined, including those with hysterectomy, endometrial ablation or contraceptive-induced amenorrhoea.

There is a need for a consistent, and implementable approach for the application, and extension of STRAW+10 criteria in research settings (5, 12, 13). Therefore, the aim of this study was to develop and validate the Reproductive Ageing in Women (RAW) Questionnaire and accompanying classification Framework. The RAW Questionnaire translates STRAW+10 criteria into a self-administered questionnaire, retaining menstrual cycle characteristics as the primary classification criteria while incorporating symptom refinement and supportive hormonal confirmation where menstrual cycle characteristics are not observable. By translating established staging criteria into an applied Framework, RAW seeks to enhance inclusivity and methodological consistency in menopausal classification across research contexts.

## 2. Methods

### 2.1. Study design

This study employed a multi-phase validation design conducted between May 2022 and July 2025. The development and validation of the RAW Questionnaire comprised four phases presented in Figure 1.

**Figure 1.**
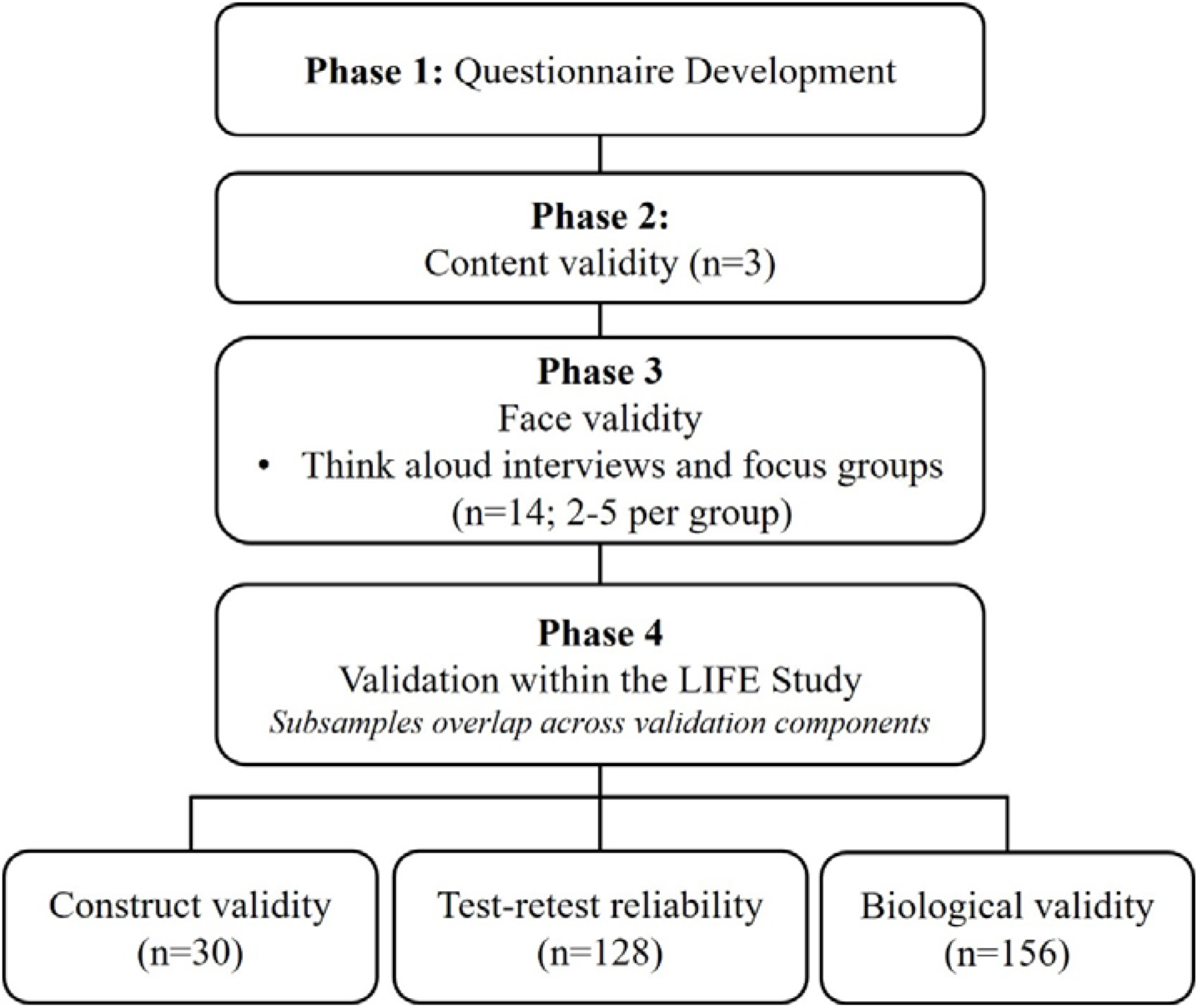
Multi-phase development and validation of the RAW Questionnaire.

### 2.2. Participant eligibility and recruitment

Participants were individuals assigned female at birth, aged ≥18 years, with current or prior menstruation recruited via convenience sampling. Exclusion criteria included pregnancy, breastfeeding, or diagnosed hormonal or ovarian disorder (e.g., polycystic ovarian syndrome, premature ovarian insufficiency). Ethical approval was granted by the University of the Sunshine Coast Human Research Ethics Committee (#S221694).

### 2.3. Phase one: Questionnaire development

Initial questions for the RAW Questionnaire were developed from STRAW+10 criteria (5), and the Menopause-Specific Quality of Life Questionnaire (MENQOL) (14) to capture menstrual cycle characteristics, symptom severity, exogenous hormone use (e.g., menopausal hormone therapy (MHT), contraceptives), and reasons for amenorrhoea. A self-classification question (premenopausal, perimenopausal, or postmenopausal), was also included.

Demographic and lifestyle variables were collected from participants completing the RAW Questionnaire during Phase three and four to describe participant characteristics. This included ethnicity, level of education, household, and marital, and employment status. Lifestyle variables included were body mass index (BMI; kg/m^2^), calculated from self-reported weight (kg) and height (cm), smoking status, and alcohol consumption (AUDIT-C).

### 2.4. Phase two: Content validity

Content validity was assessed by three menopause experts. Experts held a doctoral or medical qualification in a relevant discipline (e.g., endocrinology or epidemiology with a menopause focus) and were invited via email to review the full draft of the initial RAW Questionnaire for clarity, relevance and completeness, and usability, and provide written feedback (15). Provision of feedback was taken as implied consent to participate. Revisions were incorporated prior to subsequent phases.

### 2.5. Phase three: Face validity

Face validity was evaluated using think-aloud interviews and focus groups stratified by menopausal status. Interviews were conducted in person or online and were audio recorded. Observations from interviews were documented in real time and synthesised to inform focus group discussions. Focus group discussions explored question clarity, perceived relevance, response format, overall length, and suggestions for addition, revision or removal of questions, and were audio recorded and transcribed verbatim for thematic analysis using the transcription software (https://otter.ai).

### 2.6. Phase four: Validation within the LIfestyle risk Factors for chronic disease across the stagEs of reproductive ageing (LIFE) cohort

#### 2.6.1. LIFE study design and recruitment

Further validation of the RAW Questionnaire was completed within the LIFE study, a cross-sectional cohort study conducted at the University of the Sunshine Coast between October 2022 and May 2024. Full eligibility criteria and recruitment procedures are described in detail elsewhere (16). The RAW Questionnaire was self-administered online and completed without interviewer support prior to in-person venous blood sampling (Supplementary file 1). Ethical approval for the LIFE study was granted by the University HREC (#S211597); all participants provided written informed consent.

#### 2.6.2. Construct validity

Construct validity was assessed in 30 participants, drawn from the face validity cohort and the LIFE study. Responses were mapped against the RAW Framework to confirm that age, menstrual history, symptom severity, hormone therapy use, and surgical history could be consistently applied to yield unambiguous stage classification.

#### 2.6.3. Test re-test reliability

Test-retest reliability was assessed among LIFE participants (n=128). Percentage agreement between self-identified (single self-selection question), and RAW-classified menopausal stage was calculated. Cohen’s kappa was used for key individual classification variables (e.g., current menstrual status, cycle length, reason for cessation of menses), and Kendall’s tau-b for overall classification agreement.

#### 2.6.4. Biological validity

Venous blood samples were collected from LIFE study participants (n=156) at a single time-point in the morning in a fasted state. Premenopausal and regularly cycling perimenopausal participants, had blood drawn during the early follicular phase (days 2-7). Serum follicle stimulating hormone (FSH) and oestradiol (E2) were quantified using commercially available ELISA kits (Demeditec DE1288: Invitrogen KAQ0622) according to manufacturer guidelines and analysed in duplicate. Four parameter logistic (4PL) standard curves were generated in GraphPad Prism (version 10.6.1, GraphPad Software), and hormone concentrations were interpolated.

### 2.7. Scoring and menopausal stage classification

Menopausal stage was classified using a Framework integrating age, menstrual cycle characteristics, and hot flush severity (Figure 2). Participants with observable, naturally occurring menstrual cycles were classified according to Step 1 of the RAW Framework. Participants reporting regular endogenous cycles who did not meet RAW criteria for the menopausal transition were classified as premenopausal. Participants meeting STRAW+10 criteria for early or late menopausal transition were grouped as perimenopausal for analysis unless otherwise specified.

**Figure 2.**
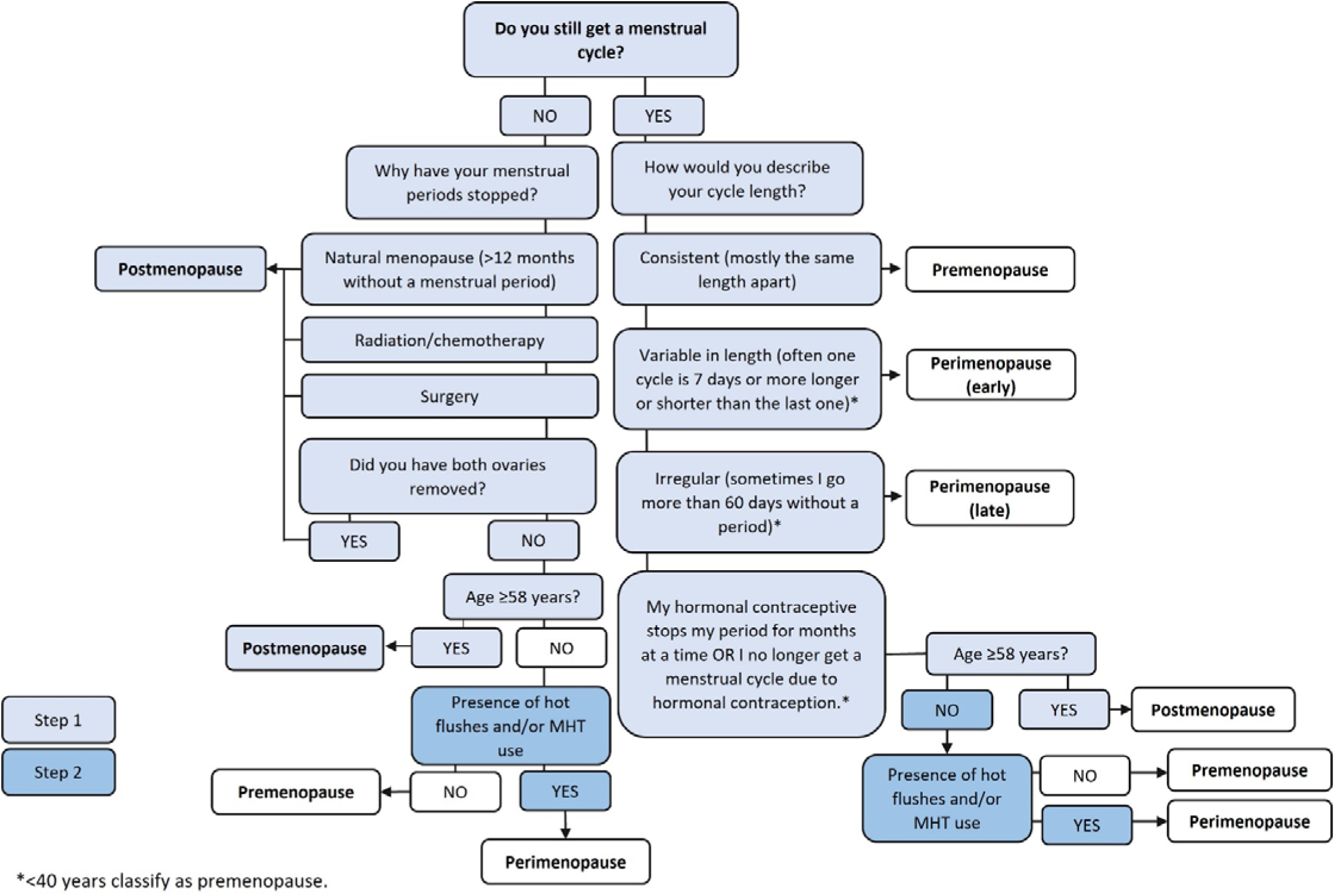
Classification Framework for the RAW Questionnaire. Classification follows sequential application of menstrual and age criteria (Step 1), and symptom refinement using moderate-to-severe hot flushes and/or MHT use (Step 2).

For participants without observable endogenous menstrual cycles (e.g., hysterectomy with ovaries retained, contraceptive induced amenorrhoea, or hormonally regulated withdrawal bleeding) were classified using a conservative age threshold (≥58 years) (Step 1), followed by refinement using moderate-to-severe hot flushes and/or MHT use (Step 2), as shown in Figure 2 (12, 17).

Hormonal contraceptive use was treated according to its likely impact on menstrual observability. Levonorgestrel-releasing intrauterine devices (IUD) users reporting ongoing regular menstrual cycles were considered to have observable cycles and were classified according to Step 1 of the RAW Framework. Where IUD use resulted in amenorrhoea, classification was based on age-based threshold in Step 1, and then proceeded to Step 2, using symptom criteria. Combined oral contraceptive users were treated as having non-observable cycles due to hormonally regulated withdrawal bleeding and were classified according to Step 1 and 2 of the Framework. Participants were assigned exclusively to one menopausal group according to the criteria outlined in Figure 2.

### 2.8. Data analysis

#### 2.8.1. Thematic analysis

Qualitative data were analysed using Braun and Clarke’s six phase thematic framework (18). Forty unique codes were identified and consolidated into five overarching themes.

#### 2.8.2. Statistical analysis

Data were analysed using SPSS (version 28.0; IBM Corp) and GraphPad Prism^®^ (version 10.6.1, GraphPad Software). Continuous variables were assessed for normality and presented as mean ± standard deviation (SD), or median and interquartile range (IQR). Categorical variables were presented as counts and proportions. Participant characteristics were grouped according to menopausal stage (pre, peri, post). Group differences were assessed using Chi-square, or Fisher’s exact tests for categorical variables, and ANOVA or Kruskal-Wallis tests with appropriate post hoc corrections for continuous variables. Significance was set at p<0.050.

For test-retest reliability, agreement in menopausal stage classified using the RAW criteria across the two time points was assessed using Kendall’s tau-b. Cohen’s kappa coefficients were calculated for individual, dichotomous classification variables (e.g. current menstrual status, exogenous hormone use, reason for cessation of menses) and interpreted according to Landis and Koch (1977) (19). Percentage agreement was calculated to assess consistency between self-report (single self-selection question), and RAW-classified menopausal stage.

## 3. Results

### 3.1. Content validity

Expert review supported the relevance and clarity of the RAW Questionnaire (15). Revisions were made to improve inclusivity and classification, including incorporation of questions adapted from the Health and Wellbeing After Breast Cancer (HWABC) menopausal classification algorithm to better capture surgical, contraceptive or medically induced amennorhoea. Clarification of unilateral versus bilateral oophorectomy and refinement of premenopausal definition were also added (12).

### 3.2. Face validity

#### 3.2.1. Participant characteristics

Fourteen participants across pre (n=6, 35.2±11.3 years), peri (n=3, 47.7±2.5 years), post (n=2, 64.0±8.5 years), and surgical induced amenorrhoea (hysterectomy with or without bilateral oophorectomy) (n=3, 52.0±8.7 years) groups completed both think-aloud interviews and focus group discussions. The majority of participants (71%) identified as Australian and held a bachelor’s degree or higher (57%).

Thematic analysis identified five themes: (1) clarity of terminology, (2) menstrual cycle descriptors, (3) menopausal symptoms, (4) usability, and (5) perceived relevance, presented in Table 1. Participant feedback informed refinement of definitions for self-identified menopausal classification, expansion of menstrual cycle descriptors, and inclusion of additional symptoms (hair loss, abdominal weight gain, difficulty to lose weight) and therapy use (e.g., over-the-counter medications, acupuncture, yoga).

**Table 1.**
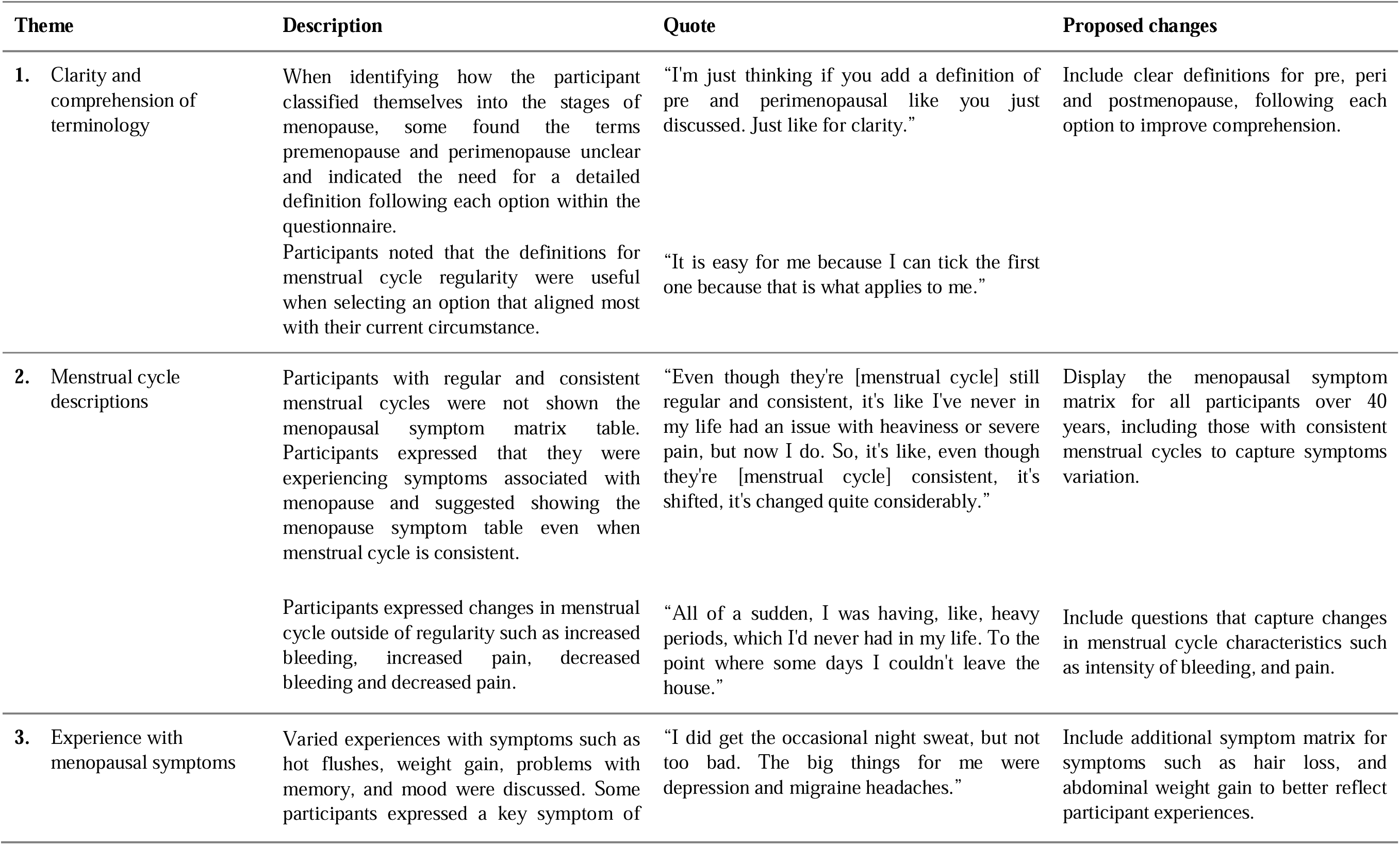
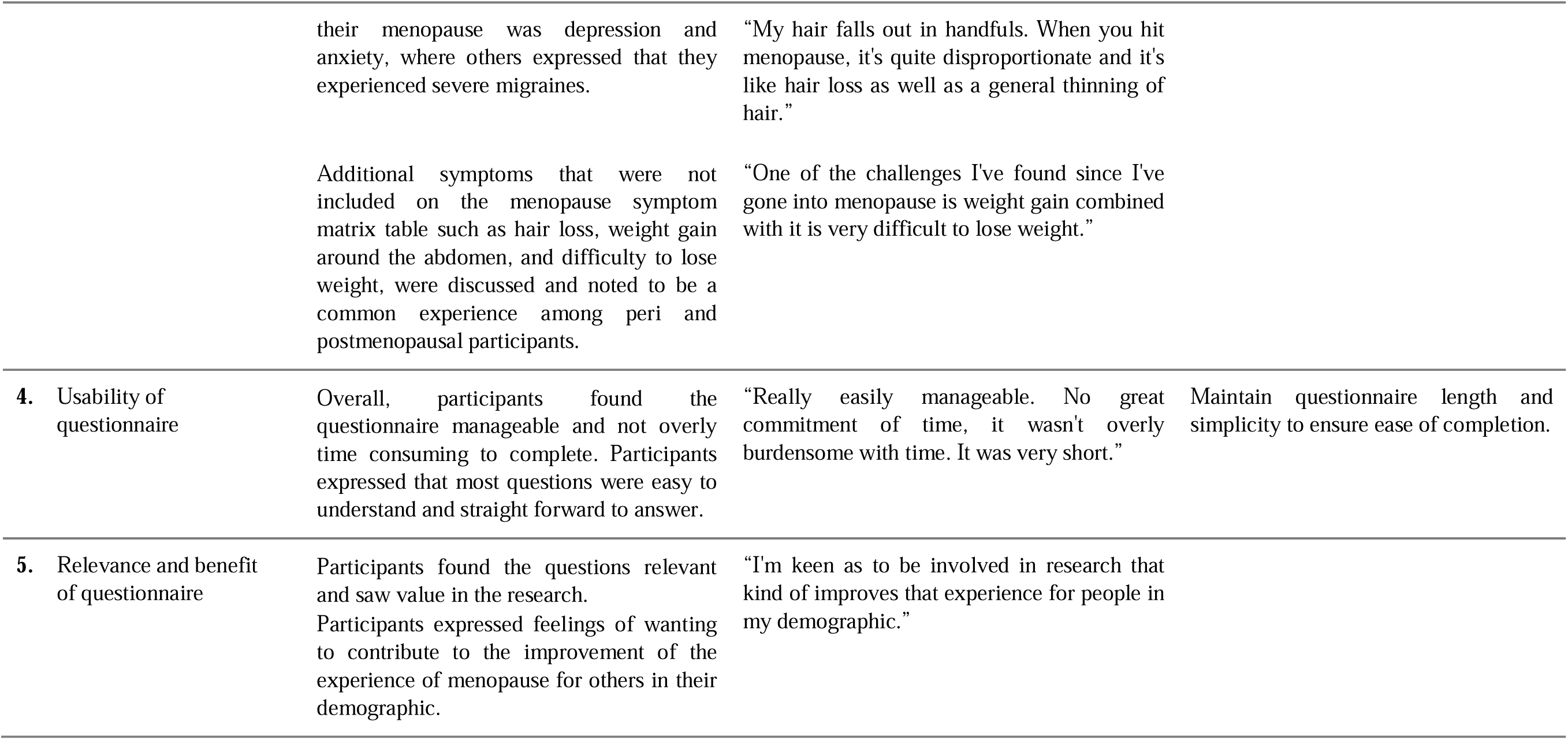
Summary of thematic analysis from face-to-face interviews and focus groups.

### 3.3. Construct validity

Construct validity was assessed in 30 participants across pre (n=11, 36.9±10.5 years), peri (n=7, 50.7±3.7 years), and postmenopausal (n=12, 58.8±5.8 years) groups. Within the perimenopausal group, two participants met criteria consistent with late perimenopause and one with early perimenopause. Early and late sub staging could not be determined for the remaining four perimenopausal participants due to the absence of an observable menstrual cycle.

Application of Step 1 (Figure 2) resulted in initial classification based on menstrual history and age. Symptom adjustment (Step 2) reclassified four participants with surgical or contraceptive-induced amenorrhoea from premenopausal to perimenopausal due to reported moderate-to-severe hot flushes. Most peri and postmenopausal participants were not using hormonal replacement therapy, however, 47.4% reported using natural therapies (e.g., vitamins, acupuncture) to manage symptoms. Moderate-to-severe hot flushes were reported by 68.6% of peri and postmenopausal participants.

RAW classification demonstrated 93% concordance with participant self-identified menopausal status. Discordance occurred in two cases. One participant self-identified as postmenopausal, however was classified as perimenopausal following Step 1 of the RAW Framework (age <58 years with surgical-amenorrhoea and moderate-to-severe hot flushes), and one self-identified as perimenopausal but met Step 1 criteria for surgical-induced amenorrhoea (age ≥58 years) and was therefore classified as postmenopausal (14).

### 3.4. Test re-test reliability

RAW classification demonstrated excellent test-retest agreement over 6-21 days (τ_b_=0.940, p< 0.001), with 95.3% agreement across two administrations in n=128 participants. Individual questions key to classification criteria demonstrated moderate to almost perfect agreement (Table 2). Although the kappa coefficient for the question ‘reason for cessation of menses’, was moderate, raw percentage agreement was high (90.6%), with discrepancies confined to a small number of responses within low frequency categories (Supplementary Table S1-S4).

**Table 2.**
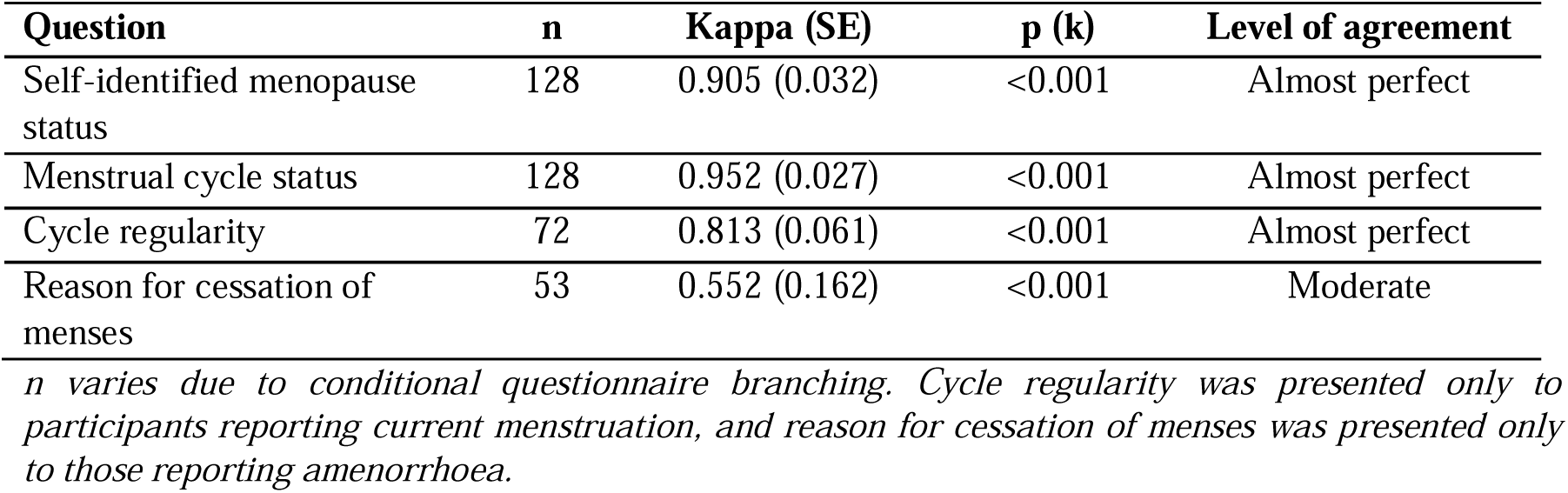
Test-retest reliability of self-identified menopausal status and key RAW classification questions across two administrations.

### 3.5. Biological validity

Biological validity (FSH and E2) was assessed in 156 participants (16). As expected, age, moderate-to-severe hot flushes, FSH and E2 levels differed significantly across stages (p<0.001). Education and alcohol intake also varied across menopausal groups (p<0.050), whereas marital status, smoking status, and BMI did not. Ethnicity and employment were not formally compared due to non-mutually exclusive response options. Detailed characteristics are presented in Table 3.

**Table 3.**
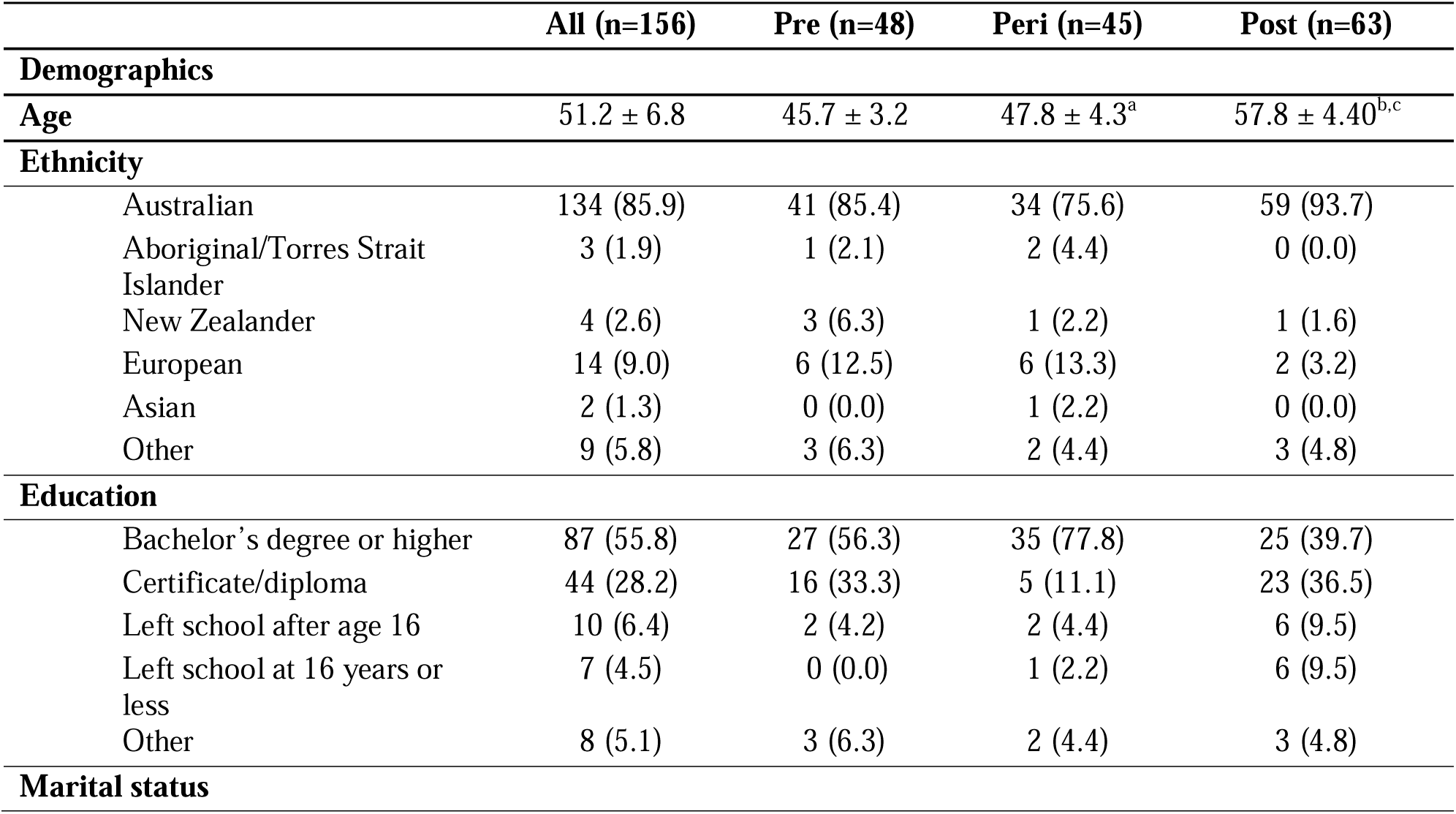
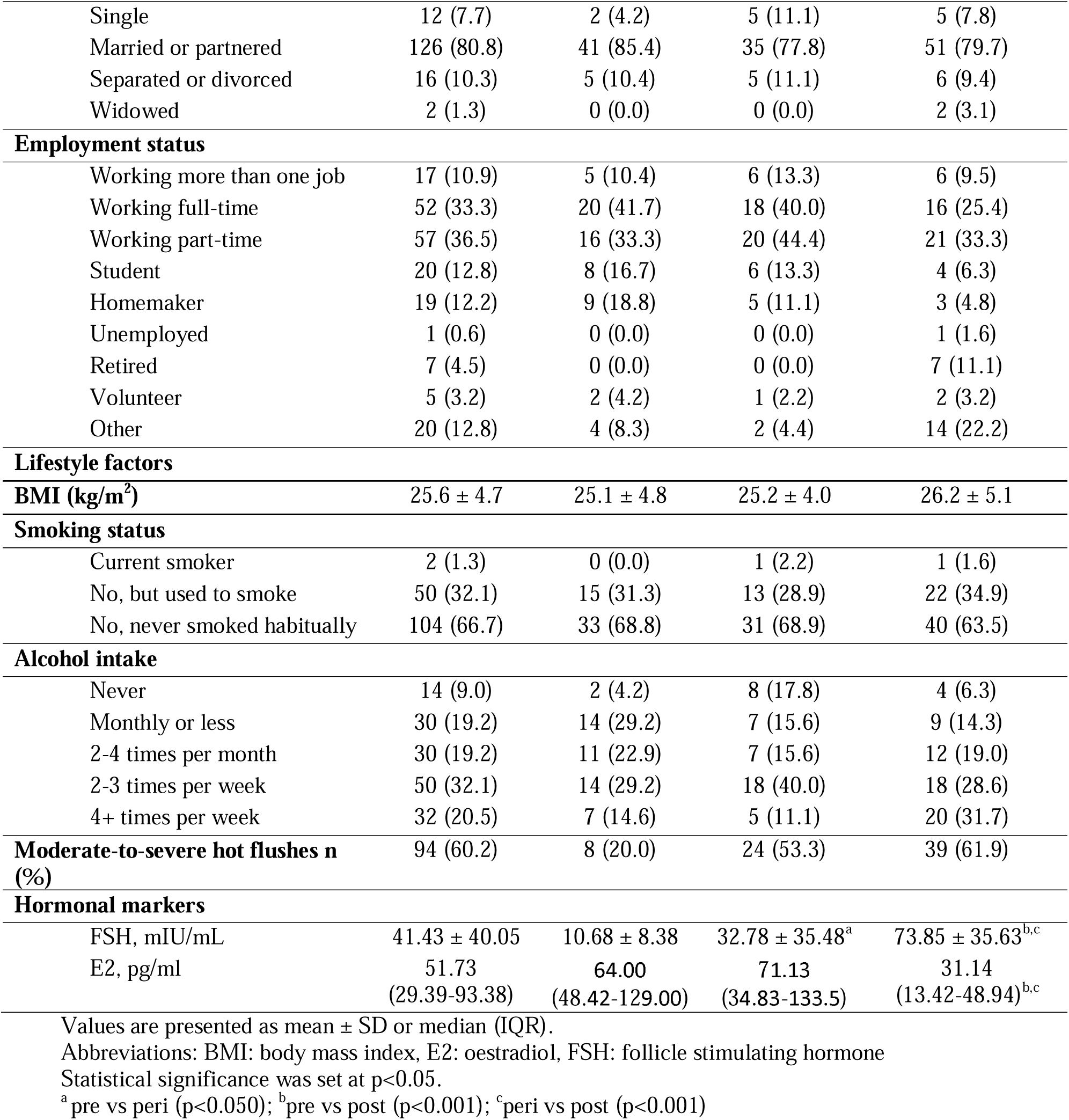
Characteristics of LIFE participants with available hormonal data, classified by menopausal stage using the RAW Framework.

|  | All (n=156) | Pre (n=48) | Peri (n=45) | Post (n=63) |
| --- | --- | --- | --- | --- |
| **Demographics** |  |  |  |  |
| Age | 51.2 ± 6.8 | 45.7 ± 3.2 | 47.8 ± 4.3<sup>a</sup> | 57.8 ± 4.40<sup>b,c</sup> |
| **Ethnicity** |  |  |  |  |
| Australian | 134 (85.9) | 41 (85.4) | 34 (75.6) | 59 (93.7) |
| Aboriginal/Torres Strait Islander | 3 (1.9) | 1 (2.1) | 2 (4.4) | 0 (0.0) |
| New Zealander | 4 (2.6) | 3 (6.3) | 1 (2.2) | 1 (1.6) |
| European | 14 (9.0) | 6 (12.5) | 6 (13.3) | 2 (3.2) |
| Asian | 2 (1.3) | 0 (0.0) | 1 (2.2) | 0 (0.0) |
| Other | 9 (5.8) | 3 (6.3) | 2 (4.4) | 3 (4.8) |
| **Education** |  |  |  |  |
| Bachelor's degree or higher | 87 (55.8) | 27 (56.3) | 35 (77.8) | 25 (39.7) |
| Certificate/diploma | 44 (28.2) | 16 (33.3) | 5 (11.1) | 23 (36.5) |
| Left school after age 16 | 10 (6.4) | 2 (4.2) | 2 (4.4) | 6 (9.5) |
| Left school at 16 years or less | 7 (4.5) | 0 (0.0) | 1 (2.2) | 6 (9.5) |
| Other | 8 (5.1) | 3 (6.3) | 2 (4.4) | 3 (4.8) |
| **Marital status** |  |  |  |  |
| Single | 12 (7.7) | 2 (4.2) | 5 (11.1) | 5 (7.8) |
| Married or partnered | 126 (80.8) | 41 (85.4) | 35 (77.8) | 51 (79.7) |
| Separated or divorced | 16 (10.3) | 5 (10.4) | 5 (11.1) | 6 (9.4) |
| Widowed | 2 (1.3) | 0 (0.0) | 0 (0.0) | 2 (3.1) |
| **Employment status** |  |  |  |  |
| Working more than one job | 17 (10.9) | 5 (10.4) | 6 (13.3) | 6 (9.5) |
| Working full-time | 52 (33.3) | 20 (41.7) | 18 (40.0) | 16 (25.4) |
| Working part-time | 57 (36.5) | 16 (33.3) | 20 (44.4) | 21 (33.3) |
| Student | 20 (12.8) | 8 (16.7) | 6 (13.3) | 4 (6.3) |
| Homemaker | 19 (12.2) | 9 (18.8) | 5 (11.1) | 3 (4.8) |
| Unemployed | 1 (0.6) | 0 (0.0) | 0 (0.0) | 1 (1.6) |
| Retired | 7 (4.5) | 0 (0.0) | 0 (0.0) | 7 (11.1) |
| Volunteer | 5 (3.2) | 2 (4.2) | 1 (2.2) | 2 (3.2) |
| Other | 20 (12.8) | 4 (8.3) | 2 (4.4) | 14 (22.2) |
| **Lifestyle factors** |  |  |  |  |
| **BMI (kg/m<sup>2</sup>)** | 25.6 ± 4.7 | 25.1 ± 4.8 | 25.2 ± 4.0 | 26.2 ± 5.1 |
| **Smoking status** |  |  |  |  |
| Current smoker | 2 (1.3) | 0 (0.0) | 1 (2.2) | 1 (1.6) |
| No, but used to smoke | 50 (32.1) | 15 (31.3) | 13 (28.9) | 22 (34.9) |
| No, never smoked habitually | 104 (66.7) | 33 (68.8) | 31 (68.9) | 40 (63.5) |
| **Alcohol intake** |  |  |  |  |
| Never | 14 (9.0) | 2 (4.2) | 8 (17.8) | 4 (6.3) |
| Monthly or less | 30 (19.2) | 14 (29.2) | 7 (15.6) | 9 (14.3) |
| 2-4 times per month | 30 (19.2) | 11 (22.9) | 7 (15.6) | 12 (19.0) |
| 2-3 times per week | 50 (32.1) | 14 (29.2) | 18 (40.0) | 18 (28.6) |
| 4+ times per week | 32 (20.5) | 7 (14.6) | 5 (11.1) | 20 (31.7) |
| **Moderate-to-severe hot flushes n (%)** | 94 (60.2) | 8 (20.0) | 24 (53.3) | 39 (61.9) |
| **Hormonal markers** |  |  |  |  |
| FSH, mIU/mL | 41.43 ± 40.05 | 10.68 ± 8.38 | 32.78 ± 35.48<sup>a</sup> | 73.85 ± 35.63<sup>b,c</sup> |
| E2, pg/ml | 51.73<br>(29.39-93.38) | 64.00<br>(48.42-129.00) | 71.13<br>(34.83-133.5) | 31.14<br>(13.42-48.94)<sup>b,c</sup> |

Values are presented as mean ± SD or median (IQR).
Abbreviations: BMI: body mass index, E2: oestradiol, FSH: follicle stimulating hormone
Statistical significance was set at p<0.05.
<sup>a</sup>pre vs peri (p<0.050); <sup>b</sup>pre vs post (p<0.001); <sup>c</sup>peri vs post (p<0.001)

Based on menstrual cycle characteristics (Step 1), 50 participants were classified as premenopausal, 43 as perimenopausal and 63 as postmenopausal. Incorporation of moderate-to-severe hot flushes (Step 2) was applied only in cases where menstrual cycle data were non-observable, resulting in reclassification of two participants from premenopausal to perimenopausal. Of the 43 perimenopausal participants with an observable menstrual cycle, 20 met criteria consistent with early perimenopause, and 23 with late perimenopause. Agreement between self-identified menopausal status and final RAW classification was 87.8%, with discrepancies occurring between pre and perimenopausal groups. In cross-sectional comparison across RAW-classified stages, the prevalence of moderate-to-s.evere hot flushes increased across menopausal stage (χ^2^ (2)=21.29, p<0.001).

Follicle stimulating hormone was significantly different across RAW-classified stages (p<0.001). Although FSH levels were not used as primary classification criteria, established reference thresholds (≤20 mIU/mL for premenopause and ≥25 mIU/mL for postmenopause) were applied to assess biological validity (5). RAW-classified pre and postmenopausal groups demonstrated 96.1% alignment with corresponding FSH thresholds. Three premenopausal participants demonstrated FSH levels above the reference threshold, two of which self-identified as perimenopausal, although menstrual criteria remained consistent with premenopause at the time of assessment. Of those classified as early perimenopause, 85% (n=17) demonstrated FSH levels ≤20 mIU/mL, and 10% (n=2) ≥25 mIU/mL. Among late perimenopausal participants, 34.8% (n=8) demonstrated FSH levels ≤20 mIU/mL, and 56.5% (n=13) ≥25 mIU/mL.

Oestradiol demonstrated expected group-level differences, with lower median levels observed in postmenopause and marked inter-individual variability (Figure 3). Incorporation of hormone data did not alter final classification when applied after menstrual and symptom criteria.

**Figure 3.**
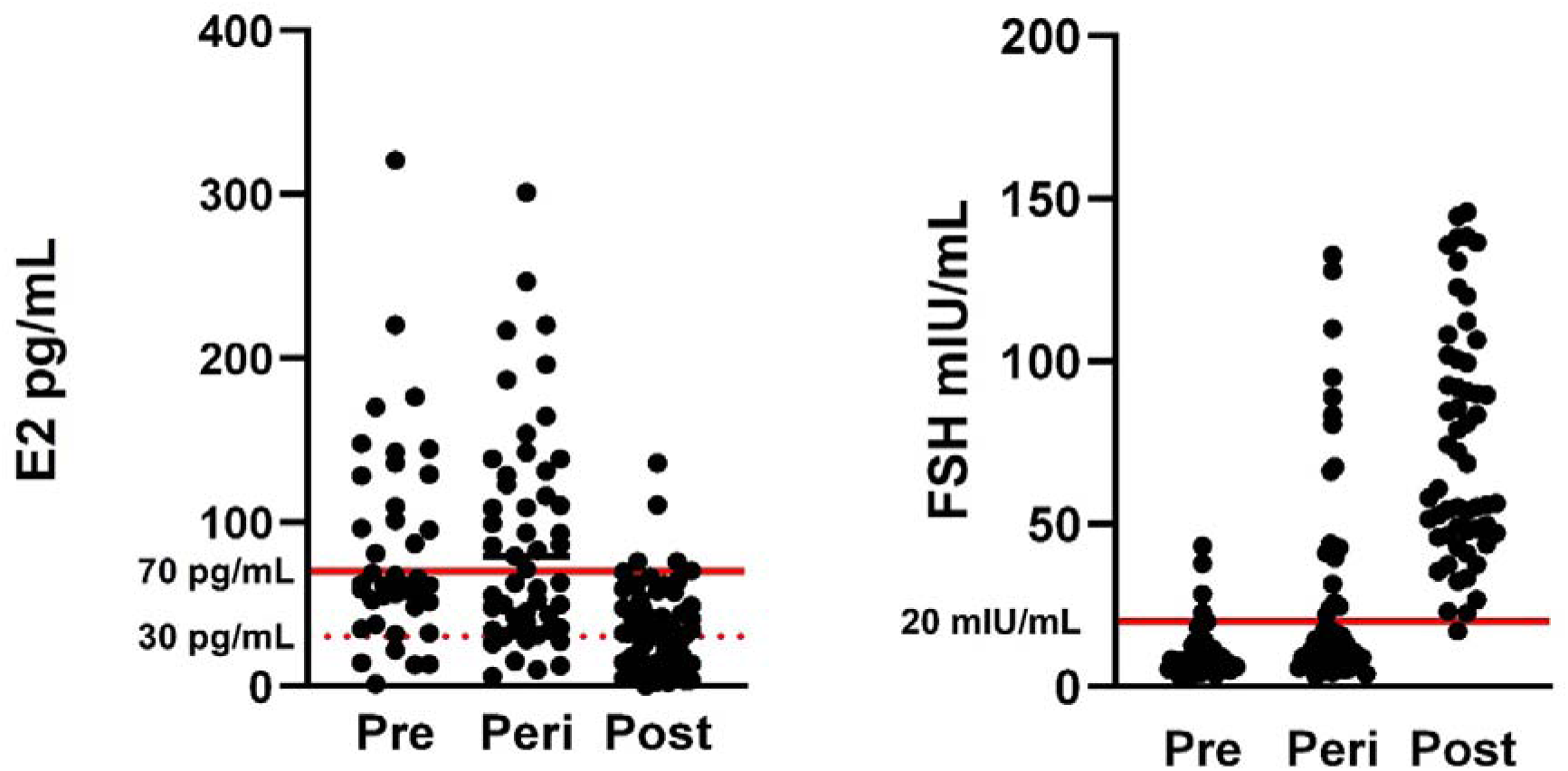
Individual serum E2 (pg/mL) and FSH (mIU/mL) levels across pre, peri and postmenopausal participants classified according to the RAW Framework. Horizontal lines at 30 and 70 pg/mL (E2) (20), and 20 mIU/mL (FSH) indicate reference thresholds (5).

## 4. Discussion

This study developed and validated the RAW Questionnaire, and Framework for menopausal stage classification in research settings. RAW demonstrated high concordance with self-identified menopausal status, short-term stability, and biological alignment with established FSH thresholds. By translating STRAW+10 into a practical Framework and extending application to individuals without observable menstrual cycles, RAW provides a standardised approach to support comparability and representation in menopausal research.

Appropriate classification of menopausal stage is fundamental to high-quality research in midlife. Although STRAW+10 provides necessary criteria for staging reproductive ageing, its operational reliance on menstrual cycle characteristics limits its applicability for individuals who do not have observable menstrual cycle patterns (5, 12). In contemporary cohorts, a considerable proportion of midlife individuals lack observable menstrual cycles due to surgical- (e.g., hysterectomy) (21) or contraceptive-induced amenorrhoea (22). Consequently, individuals without observable cycles are often excluded from population-level menopausal research (2). Restricting analyses to those with evaluable bleeding patterns narrows the study population to a subset of midlife individuals whose reproductive ageing follows an observable trajectory. To address this limitation where menstrual data are unavailable, RAW applies criteria, including conservative age thresholds and hot flush severity, to enable pragmatic classification in cross-sectional research. Algorithm-based approaches have previously incorporated intervention history and age thresholds to improve menopausal classification in large epidemiological cohorts (17). While such approaches enhance classification where menstrual data are incomplete, they are typically designed to optimise exposure classification for disease modelling rather than to evaluate stage-specific physiological characteristics. RAW aligns with this methodological direction and extends it by incorporating symptom refinement and/or MHT use within a structured Framework applicable to cross-sectional studies.

Although STRAW+10 does not incorporate vasomotor symptoms into primary staging criteria (5), emerging evidence suggests that moderately-to-severely bothersome vasomotor symptoms may precede sustained menstrual cycle irregularity in a subset of individuals (13, 23, 24). Islam et al. (13) reported substantial increases in vasomotor symptom prevalence across the late reproductive stage and early menopausal transition, with symptomatic overlap between adjacent bleeding-defined stages, raising the possibility that thermoregulatory instability may emerge before overt cycle disruption (25).

In the present study, a small proportion of regularly cycling participants reported moderate-to-severe hot flushes. While it is plausible that some of these individuals were at the onset of the early menopausal transition, endocrine profiles did not demonstrate elevations in FSH, or lower E2 relative to expected premenopausal distribution (20). Given the recognised intra-individual variability in FSH and E2 during the late reproductive stage, symptom reports in isolation do not provide sufficient evidence to justify stage reclassification in cross-sectional design (20). However, the presence of vasomotor symptoms in regularly cycling individuals warrants longitudinal investigation. Repeated hormonal assessment and prospective menstrual tracking are required to determine whether early vasomotor symptoms in this subgroup represent transient fluctuation or the earliest endocrine manifestations of transitions preceding overt cycle disruption.

Within RAW, hormone measures were incorporated as confirmatory rather than deterministic criteria. Using reference thresholds (5), FSH demonstrated 96.1% concordance with RAW pre and postmenopausal classifications derived from menstrual cycle characteristics. However, variability across participants in perimenopause was consistent with the well-described endocrine instability of this stage (2). Isolated higher FSH levels were also observed in a small number of regularly cycling participants, without moderate-to-severe vasomotor symptoms. This aligns with prior evidence demonstrating transient FSH elevations during the late reproductive stage that do not necessarily indicate sustained ovarian insufficiency (26). In cross-sectional contexts, reliance on single time-point hormone thresholds risks over classification of menopausal transition staged and may conflate physiological variability with categorical change.

These observations reinforce a broader methodological consideration in menopausal research. Hormonal biomarkers provide valuable biological context but have limited discriminatory capacity when interpreted in isolation. Consecutive measurements may improve staging accuracy, particularly in individuals without observable menstrual cycles due to hysterectomy or non-hormonal contraception (5). However, in large-scale or cross-sectional studies where repeated sampling is not feasible, anchoring classification to menstrual cycle characteristics (5), supplemented by conservative age-thresholds (12) and symptom data (13) where necessary, may more closely reflect reproductive ageing.

A central contribution of this study is the translation of STRAW+10 primary menstrual criteria into a validated questionnaire and Framework, alongside pre-specified extensions for individuals in whom menstrual cycle characteristics are not observable. Although STRAW+10 provides criteria for staging reproductive ageing, its direct application in cross-sectional research and in populations affected by surgical or contraceptive-induced amenorrhoea is inherently constrained. By delineating how classification proceeds both when menstrual criteria are observable and when they are not, the RAW Framework addresses a methodological gap between staging principles and their practical implementation in contemporary cohort research.

Several limitations warrant consideration. Classification in the absence of menstrual cycles remains inherently challenging (5, 12, 13). A single conservative age-based threshold (12) was applied rather than procedure-specific thresholds for ease of use, however cross-sectional data cannot fully compensate for missing menstrual or follow-up hormone data. In addition, RAW is not appropriate for individuals with POI, as classification is anchored to normative, age-related reproductive ageing patterns (5). Use of exogenous hormone may obscure endogenous reproductive status. Combined oral contraceptives can regulate withdrawal bleeding and may attenuate vasomotor symptoms. Although the RAW Framework accounted for hormonal contraception by treating these cycles as non-observable, attenuation of symptom reporting may have reduced the sensitivity of symptom-based refinement, introducing the possibility of misclassification (27).

Finally, the cross-sectional design precludes evaluation of temporal sensitivity. Menopause is a dynamic process characterised by hormonal and symptomatic fluctuation (28). Although short-term test-retest reliability was high, longitudinal validation is needed to determine whether RAW can reliably distinguish late reproductive stage, and early menopause transition over time (2, 28, 29).

## 5. Conclusions

This study developed and validated the RAW Questionnaire and accompanying Framework for menopausal stage classification in research settings. Through application of STRAW+10 menstrual criteria and defining a conservative approach for individuals without observable menstrual cycles, RAW provides a practical approach for classifying menopausal stage. Menstrual cycle characteristics remained the primary determinant for classification, with symptom and hormone data incorporated where indicated. As midlife is increasingly recognised as a critical window for disease risk modification, consistent and inclusive staging frameworks are essential to ensure valid stage-specific analyses and comparability across studies. Further longitudinal validation is required to assess temporal sensitivity and refine the integration of symptom and hormone data across the menopausal transition.

## Supporting information

Supplementary material_RAW Questionnaire

## Data Availability

All data produced in the present study are available upon reasonable request to the authors.

## Funding

This work was supported by the University of the Sunshine Coast LAUNCH Grant (grant number 980027659) awarded to M.S, M.D, and Alexandra Metse. L.P is supported by the University of the Sunshine Coast, Deputy Vice Chancellor of Research, and Innovation scholarship, J.N was supported by the University of the Sunshine Coast, Higher Degree by Research Program Support Grant.

## Acknowledgements

The authors would like to thank all the content experts, and participants who volunteered their time to partake in this study and Dr Alexandra Metse for her contribution to securing study funding.

## Notes

### Competing Interest Statement

The authors have declared no competing interest.

### Funding Statement

This study was funded by the University of the Sunshine Coast LAUNCH Grant (grant number 980027659) awarded to M.S, M.D, and Alexandra Metse. L.P is supported by the University of the Sunshine Coast, Deputy Vice Chancellor of Research, and Innovation scholarship, J.N was supported by the University of the Sunshine Coast, Higher Degree by Research Program Support Grant.

### Author Declarations

Human research ethics committee of the University of the Sunshine Coast gave ethical approval for this work.

