## Supplementary material_RAW Questionnaire for "Reproductive Ageing in Women (RAW) Questionnaire: multi-phase development and validation of a questionnaire for the classification of menopause stage"

Copyright: Laura Pernoud & University of the Sunshine Coast 2026.

This questionnaire may be used freely for research purposes. The authors request acknowledgment in any research publications in which the questionnaire is used.

usc.edu.au

**The Reproductive Ageing in Women (RAW) Questionnaire**

**Overview**

The RAW is a self-report questionnaire with accompanying classification framework designed to determine the stages of reproductive ageing using menstrual history, vasomotor symptoms, hormone levels (where available), hormone therapy and contraception use, and surgical history (e.g., hysterectomy).

RAW applies a structured, stepwise framework, with pragmatic refinements to accommodate cases where menstrual bleeding is non-observable.

**Administration**

Exclusions: primary ovarian insufficiency, hypothalamic amenorrhea, premature menopause (<40 years), pregnancy/postpartum amenorrhoea were excluded.

Format: Self-administered, online compatible with branching logic

**Intended scope**

RAW is designed for research settings where menstrual data are self-reported and endocrine measures may not be repeatedly available.

In cases where menstrual bleeding is non-observable (e.g., endometrial ablation, hysterectomy without bilateral oophorectomy, or contraception-related amenorrhoea), classification incorporates a conservative age-based threshold (≥58 years) and symptom/MHT refinement for those <58 years. For these individuals, classification reflects probability rather than direct menstrual confirmation and should be interpreted with caution.

**SECTION A: DEMOGRAPHIC INFORMATION**

Q1. **Date:**
Q2. **Date of birth (dd/mm/yyyy):**
Q3. **What is your height (cm)?**
Q4. **What is your weight (kg)?**
Q5. **Which gender do you identify with?**

- Cisgender woman (cisgendered refers to a person whose sex assigned at birth and gender identity are the same e.g., if you were born a woman and currently identify as a women).
- Non-binary (for the purpose of this research study, non-binary includes those whose sex at birth was female, and gender identity is non-binary)
- Decline to answer
- Other, please specify

Q6. **Which ethnic group do you identify with?**

**Please select as many options as you identify with**

- Australian
- Aboriginal or Torres Strait Islander
- New Zealander
- Pacific Islander
- North-west European
- Southern and Eastern European
- North African and Middle Eastern
- South-East Asian
- North-East Asian
- Southern and Central Asian
- Peoples of the Americas
- Other, please specify

**Smoking**

Q7. **Do you currently smoke cigarettes?**

- YES

How many per day?

At what age were you when you started smoking cigarettes (yrs)?

- NO, but I used to smoke

At what age were you when you started smoking cigarettes (yrs)?

At what age did you stop (yrs)?

- NO, I have never smoked a cigarette

**Alcohol (AUDIT-C)**

Q8. **How often do you have a drink containing alcohol?**

- Never
- Monthly or less
- 2-4 times per month
- 2-3 times per week
- 4+ times per week

Q9. **How many Australian standard alcoholic drinks do you drink on a typical day**

**when you are drinking?**

- 1-2
- 3-4
- 5-6
- 7-9
- 10+

Q10. **How often have you had 6 or more Australian standard drinks containing**

**alcohol on a single occasion in the last year?**

- Never
- Less than monthly
- Monthly
- Fortnightly
- Weekly
- Daily, or almost daily

Q11. **What best describes the highest level of education you have obtained?**

- Left school at 16 years or less
- Left school after age 16
- Trade / Apprenticeship
- Certificate / Diploma
- Bachelor degree or higher
- Other, please specify ____________
- Prefer not to say

Q12. **What was your employment status over the last 3 months?**

**Were you primarily ….. (multiple response)**

- Working more than one job
- Working full-time
- Working part time
- A student
- A homemaker
- Unemployed
- Retired
- Volunteer
- Other (please specify)
- Prefer not to say
- Don’t know

Q13. **If answered a-d, thinking about the past 3 months, which of the following best**

**describes your work schedule? Would you say that you worked… (single response)**

- Regular day shifts
- Regular evening shifts
- Regular night shifts
- Rotating night shifts
- Other (please specify)
- Prefer not to say
- Don’t know

Q14. **On average, how many total hours per week do you work at a job for which you**

**are paid?**

- <10hrs
- 10-15 hrs
- 15-20 hrs
- 20-25 hrs
- 25-30 hrs
- 30-35 hrs
- 35-38 hrs
- >38 hrs

Q15. **If studying, on average how many total hours per week do you spend studying?**

- <10hrs
- 10-15 hrs
- 15-20 hrs
- 20-25 hrs
- 25-30 hrs
- 30-35 hrs
- 35-38 hrs
- >38 hrs

Q16. **If volunteering, on average, how many total hours per week do you spend**

**volunteering?**

- <10hrs
- 10-15 hrs
- 15-20 hrs
- 20-25 hrs
- 25-30 hrs
- 30-35 hrs
- 35-38 hrs
- >38 hrs

Q17. **What is your Occupation: ____________________________**
Q18. **Which of the following best describes your current relational status?**

- Single
- Married or partnered
- Separated or divorced
- Widowed
- Prefer not to say

Q19. **Who else lives in your household with you?**

- My partner/spouse
- My children
- Parent (s)
- Siblings(s) e.g., Brother, Sister
- Other, please specify: ______________
- None of the above/live alone

Q20. **If you have children, how many children do you have?**

- 1
- 2
- 3
- 4 or more

Q21. **If living at home with children, please indicate the age category of the children**

**who live with you.**

- 0-6 months (newborn)
- 7-23 months (Infant)
- 2-5 yrs (Preschool)
- 6-18 yrs (School Age)
- 19 yrs or older (adult)

**SECTION B: MENSTRUAL HISTORY**

Q22. **How old were you when you had your first menstrual period?**

- 9 or less
- 9-10
- 10-11
- 11-12
- 12-13
- 13-14
- 14-15
- 15-16
- 16 or older

Q23. **How would you describe your current menstrual status?**

- Premenopause (before menopause, not experiencing menopausal symptoms, mostly consistent menstrual cycles)
- Perimenopause/menopausal transition (onset of menopausal symptoms, change in menstrual cycles, but have not gone 12 months in a row without a period)
- Postmenopause (after menopause, have gone 12 months or more without a period)

Q24. **Do you still get your menstrual period?**

- YES
- NO (proceed to Q10)
- My contraception suppresses my menstrual period

**IF YES:**

Q25. **How would you describe your cycle length (the number of days between the start of one period and the start of the next)?** *Select the response that most applies*

- Consistent: mostly about the same length apart
- Variable in length: often one cycle is 7 days or more longer or shorter than the last one
- Irregular: I sometimes go more than 60 days without a period
- My hormonal contraceptive stops my periods for months at a time

Q26. **How long have your menstrual periods been this way**

__ Months

__ Years

Q27. **Have you noticed any changes in your menstrual bleeding pattern, including heaviness of flow, or increase/decrease in pain during menstruation?**

- YES
- NO (proceed to Q8)

Q28. **IF YES:**

**Please select all that are relevant to you**

- Subtle change in menstrual cycle regularity (<7 days longer or shorter between cycles)
- Increase/decrease in menstrual flow, please specify:
- Increase/decrease in pain during menstruation, please specify:

Q29. **Are you currently taking/on hormonal contraception?**

- YES
- NO (proceed to Section D)

Q30. **IF YES (after answering Q9, proceed to section D):**

**Please select the type of hormonal contraception:**

- Oral contraceptive pill (e.g., combination pill)
- Hormonal implant (e.g., Implanon)
- Contraceptive injection (e.g, Depot)
- Contraceptive patch (e.g., birth control patch)
- Intrauterine device (e.g., Mirena, copper IUD)
- Other, please specify: ____________________

**IF NO LONGER GET A MENSTRUAL CYCLE:**

Q31. **Why have your periods stopped?**

- Due to natural menopause
- Surgery
- Due to radiation therapy/chemotherapy.

Please specify type of radiation/chemotherapy: ______________________

- Due to other medical reasons? Please specify: _______________________
- My contraception stops my period (*please answer Q9*)
- I am not sure of the reason

Q32. **How long ago did your periods stop?**

_________ years

_________ months

Q33. **If your periods stopped due to surgery, what was the type of surgery?**

- Endometrial ablation (proceed to Section C)
- Hysterectomy (removal of the uterus) (proceed to Section C)
- Oophorectomy (removal of one or both ovaries)
- Both, hysterectomy, and oophorectomy
- Other, please specify: __________________

Q34. **IF you have had both your uterus and ovaries removed, was this surgery completed at the same time?**

	YES (Proceed to Section C)

	NO

**14. IF NO:**

How many years ago was your hysterectomy? (Removal of uterus) ___________

How many years ago was your oophorectomy? (Removal of ovaries) __________

**SECTION C: MENOPAUSE TREATMENT**

Q35. **Do you currently use menopausal hormone therapy?**

- YES (Go to Q38)
- NO

Q36. **IF NO:**

**Have you used hormonal therapy for menopause in the past?**

- YES
- NO

Q37. **IF YES:**

How long ago did you stop the hormonal therapy? ________

How long were you taking hormonal therapy for (approx.)? ______________

Q38. **Have you taken/or are taking any medications and/or supplements (including vitamins, calcium, herbs, soy, melatonin etc.) to treat menopausal related symptoms (e.g., Sleep, weight gain, anxiety etc.)?**

- YES
- NO (proceed to Q24)

Q39. **IF YES:**

*Please indicate any medications and/or supplements*

- Soy
- Calcium
- Melatonin
- Vitamins, please specify: _________________
- Herbs, please specify: ____________________
- Other, please specify: ____________________

Q40. **How long have/were you been taking this supplement for?**

____ months

____ years

Q41. **Are you still taking this supplement?**
     - YES
     - NO
Q42. **IF NO:**

How long ago did you stop taking this supplement?

______ months

______ years

Q43. **Have you found this therapy to be helpful in managing these symptoms?**

- YES
- NO
- Unsure

Q44. **Have you used any other therapy (besides medication/supplements) for menopause-associated symptoms (such as acupuncture or yoga)?**

- YES
- NO

Q45. **IF YES**

**Have you found this therapy to be helpful in managing these symptoms?**

- YES
- NO (Go to Q47)
- Unsure (Go to Q47)

Q46. **IF YES**

**Which therapy have you found helpful for managing your menopause-associated symptoms?**

- Acupuncture
- Yoga
- Herbs
- Vitamins
- Melatonin
- Calcium
- Soy
- Other, please specify
- I am unsure which therapy has been helpful in managing my menopause
- symptoms

Q47. **Apart from the medications already indicated, do you take any other prescribed**

**medication for other health conditions e.g., (Depression, osteoarthritis, bone loss etc.)**

- YES, please specify: __________________
- NO

**SECTION D: MENOPAUSE SYMPTOMS**

Menopausal symptoms were assessed using the validated Menopause-Specific Quality of Life (MENQOL) questionnaire. The instrument was administered in its original format, and hot flush severity (rated as ≥3) was used to inform symptom-based refinement within the RAW framework.

In addition to the validated MENQOL items, participants were asked to report additional commonly described menopausal symptoms for descriptive purposes.

Q48. **For each of the following items, indicate whether you have experienced the**

**problem in the PAST MONTH.**

**If you have, rate how much you have been *bothered* by the problems**

|  | Not at all bothered | | 0 | 1 | 2 | 3 | 4 | 5 | 6 | Extremely bothered |
| --- | --- | --- | --- | --- | --- | --- | --- | --- | --- | --- |
| Weight gains around the abdomen |   No |   Yes | 🡪 | 0 | 1 | 2 | 3 | 4 | 5 | 6 |
| Difficulty to lose weight |   No |   Yes | 🡪 | 0 | 1 | 2 | 3 | 4 | 5 | 6 |
| Hair loss |   No |   Yes | 🡪 | 0 | 1 | 2 | 3 | 4 | 5 | 6 |

22. **Please note any additional symptoms you may be experiencing due to menopause:**

**RAW STEPWISE CLASSIFICATION FRAMEWORK**

**STEP 1: MENSTRUAL CYCLE CHARACTERISTICS**

1. **Observable cycles**
   a. Regular cycles 🡪 Premenopause (provisional; subject to step 2 refinement)
   b. ≥7-day variability between consecutive cycles 🡪 Early perimenopause
   c. ≥60 days between menstrual cycles (<12 months total) 🡪 Late perimenopause
2. **Natural amenorrhoea**
a. ≥12 consecutive months without menstruation 🡪 Postmenopause
3. **Surgical menopause**
   a. Bilateral oophorectomy ± hysterectomy 🡪 Postmenopause
4. **Non-observable cycles**
   a. Age ≥58 years 🡪 Postmenopause
   b. Age <58 years 🡪 Premenopause (provisional; subject to symptom refinement)

**STEP 2: VASOMOTOR SYMPTOM RECLASSIFICATION**

Applied only to individuals aged ≥40 years with non-observable cycles (<58 years)

**IF:**

1. Hot flush severity score ≥3(MENQOL)

**OR**

Hormone replacement therapy (HRT) use 🡪 Reclassify to perimenopause

2. Hot flush severity score <3 and no HRT use (MENQOL) 🡪 Remain premenopause

**NOTE:** Symptom and HRT refinement did not override classifications determined by bilateral oophorectomy, ≥12 months natural amenorrhoea, or age ≥58 in non-observable cycles.

FSH was used as supportive confirmation of stage where available but did not override the above criteria.

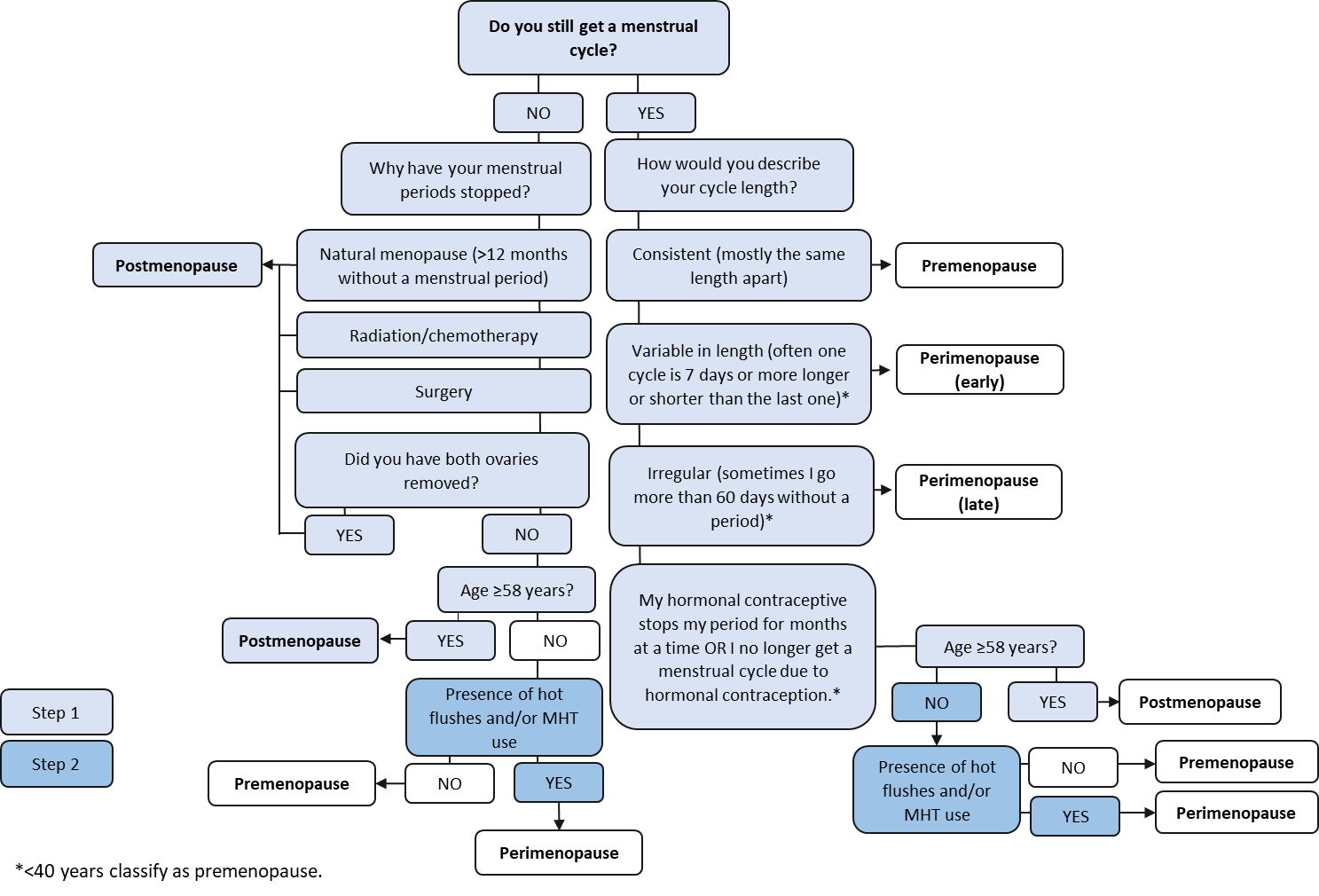
