## Supplementary material for "Reproductive Ageing in Women (RAW) Questionnaire: multi-phase development and validation of a questionnaire for the classification of menopause stage"

**Journal: Maturitas**

**SUPPLEMENTARY MATERIAL**

**Supplementary file 1: The Reproductive Ageing in Women Questionnaire (RAW)**

**Supplementary table 1. Cross-tabulation of RAW-classified menopausal stage classification at Time 1 and Time 2 (n=128) for self-identified menopausal status: *How would you describe your current menstrual status?***

**Supplementary table 2. Cross-tabulation of RAW-classified menopausal stage classification at Time 1 and Time 2 (n=128) for menstrual cycle status: *Do you still get your menstrual cycle?***

**Supplementary table 3. Cross-tabulation of RAW-classified menopausal stage classification at Time 1 and Time 2 (n=128) for cycle regularity: *How would you describe your cycle length?***

**Supplementary table 4. Cross-tabulation of RAW-classified menopausal stage classification at Time 1 and Time 2 (n=128) for reason for cessation of menstrual period: *Why have your periods stopped?***

**Supplementary table 1. Cross-tabulation of RAW-classified menopausal stage classification at Time 1 and Time 2 (n=128) for self-identified menopausal status: *How would you describe your current menstrual status?***

|  | Pre | Peri | Post | Total |
| --- | --- | --- | --- | --- |
| Pre | 31 | 2 | 1 | 34 |
| Peri | 5 | 40 | 0 | 45 |
| Post | 0 | 0 | 49 | 49 |
| Total | 36 | 42 | 50 | 128 |

**Supplementary table 2. Cross-tabulation of RAW-classified menopausal stage classification at Time 1 and Time 2 (n=128) for menstrual cycle status: *Do you still get your menstrual cycle?***

|  | Yes | No | Total |
| --- | --- | --- | --- |
| Yes | 72 | 2 | 74 |
| No | 1 | 53 | 54 |
| Total | 73 | 55 | 128 |

**Supplementary table 3. Cross-tabulation of RAW-classified menopausal stage classification at Time 1 and Time 2 (n=128) for cycle regularity: *How would you describe your cycle length?***

|  | Regular | Variable | Irregular | Total |
| --- | --- | --- | --- | --- |
| Regular | 37 | 1 | 1 | 39 |
| Variable | 4 | 12 | 2 | 18 |
| Irregular | 0 | 0 | 15 | 15 |
| Total | 41 | 13 | 18 | 72 |

**Supplementary table 4. Cross-tabulation of RAW-classified menopausal stage classification at Time 1 and Time 2 (n=128) for reason for cessation of menstrual period: *Why have your periods stopped?***

|  | Natural menopause | Surgery | Radiation/  chemotherapy | My hormonal contraception stops my period | I am not sure of the reason | Total |
| --- | --- | --- | --- | --- | --- | --- |
| Natural menopause | 45 | 1 | 0 | 1 | 0 | 47 |
| Surgery | 0 | 2 | 0 | 0 | 0 | 2 |
| Radiation/  chemotherapy | 0 | 0 | 1 | 0 | 0 | 1 |
| Due to other medical reasons | 1 | 0 | 0 | 0 | 1 | 2 |
| My hormonal contraception stops my period | 1 | 0 | 0 | 0 | 0 | 1 |
| Total | 47 | 3 | 1 | 1 | 1 | 53 |
